# Pathway-specific population attributable fractions

**DOI:** 10.1101/2020.10.15.20212845

**Authors:** Maurice O’Connell, John Ferguson

## Abstract

A population attributable fraction (PAF) represents the relative change in disease prevalence that one might expect if a particular exposure was absent from the population. Often, one might be interested in what percentage of this effect acts through particular pathways. For instance, the effect of excessive alcohol intake on stroke risk may be mediated by blood pressure, body mass index and several other intermediate risk factors. In this situation, attributable fractions for each mediating pathway of interest can be defined as the relative change in disease prevalence from disabling the effect of the exposure through that mediating pathway.

This quantity is related to, but distinct from the recently proposed metrics of direct and indirect PAF by Sjölander. In particular, while differing pathway-specific PAF will each usually be less than total PAF, they may sum over differing mediating pathways to more than total PAF, whereas direct and indirect PAF must sum to total PAF. Here, we present definitions, identifiability conditions and estimation approaches for pathway-specific attributable fractions. We illustrate results, and comparisons to indirect PAF using INTERSTROKE, a case-control study designed to quantify disease burden attributable to a number of known causal risk factors.

## Introduction

Population attributable fractions (PAF) represent the relative change in disease prevalence that one might expect if a particular exposure was absent from the population. This metric was originally introduced by [1] in 1952 to estimate the percentage of lung cancer that would not have occurred under a counterfactual scenario that nobody smoked from the population. Since then, these family of metrics have become a standard way of measuring total disease burden attributable to a risk factor [2], and also to rank differing risk factors for prioritization as intervention targets [3].

Partitioning this overall disease burden into contributions from the known pathways through which the risk factor affects disease is also useful both in understanding pathogenic mechanisms and also when comparing interventions that may reduce disease. For instance, we might estimate that in a hypothetical world, where dietary red-meat was completely substituted for plant based protein, the prevalence of heart disease might be reduced by 10%. How much of this reduction in disease burden is attributable to the pathway by which diet affects blood pressure? Here, we introduce the Pathway Specific population attributable fraction (PS-PAF) to help answer this question. In the preceding example, the PS-PAF can be informally understood as the relative change in disease prevalence in a hypothetical world where the distribution of blood pressure was altered to match the distribution expected under the afore-mentioned dietary substitution. Under certain assumptions (described later), the same quantity can be described more mechanistically as the proportion of disease that would be avoided from completely disabling the corresponding mediating pathway (here the pathway is diet → blood pressure → heart disease). PS-PAFs can be calculated for multiple mediating pathways for the same exposure. For example, the effect of the previous dietary substitution might be partially mediated by the effect on cholesterol as well as blood pressure; separate PS-PAFs could be compared for both pathways. In this case, the aim is not to provide a decomposition of the overall attributable fraction into the mediating pathways, and we argue later that such a decomposition would not be sensible. Just as differing attributable fractions for a set of risk factors typically sum to more than the joint attributable fraction [15], differing PS-PAFs corresponding to various mediating pathways between a particular risk factor and disease typically sum to more than the overall PAF for the risk factor. Rather than decomposing the total PAF, the aim instead is to fairly compare disease burden attributable to differing pathways and as a result gain insights into the dom-inant mechanisms by which the risk factor affects disease on a population level. These insights may in turn be useful in comparing possible interventions to prevent disease.

In the next Section, we will introduce notation and carefully describe PS-PAFs under various identifiability assumptions. We also will also compare and contrast PS-PAFs with the related concepts of indirect and direct PAF recently introduced by Sjölander [13]. Later, we illustrate results using data from INTERSTROKE, a case-control study designed to quantify disease burden attributable to a number of known causal risk factors for stroke. A further discussion section concludes the manuscript.

## Methods

### Potential outcome notation used for mediation analyses

We borrow notation from Vanderweele, [16], in defining nested counterfactuals. As is usual in the causal inference literature, random variables for observed variables will be denoted with unscripted notation, and potential outcomes will be denoted using subscripts. In all cases, we use upper case letters to denote random quantities, and lower-case to denote quantities that are fixed or intervened on. In particular, let *C* denote a vector of known baseline covariates not effected by the exposure, *A* ∈ {0, 1*}* a binary exposure of interest and *M* ^1^, …*M*^*K*^ mediatos on separate causal pathways from *A* to *Y*, (note that each *M*^*k*^ could be binary, multi-category or continuous). Finally *Y* ∈ {0, 1*}* is a binary disease outcome. Figure 1 below demonstrates a multi-mediator scenario with 3 mediators, *M* ^1^, *M* ^2^ and *M* ^3^.

**Figure 1:**
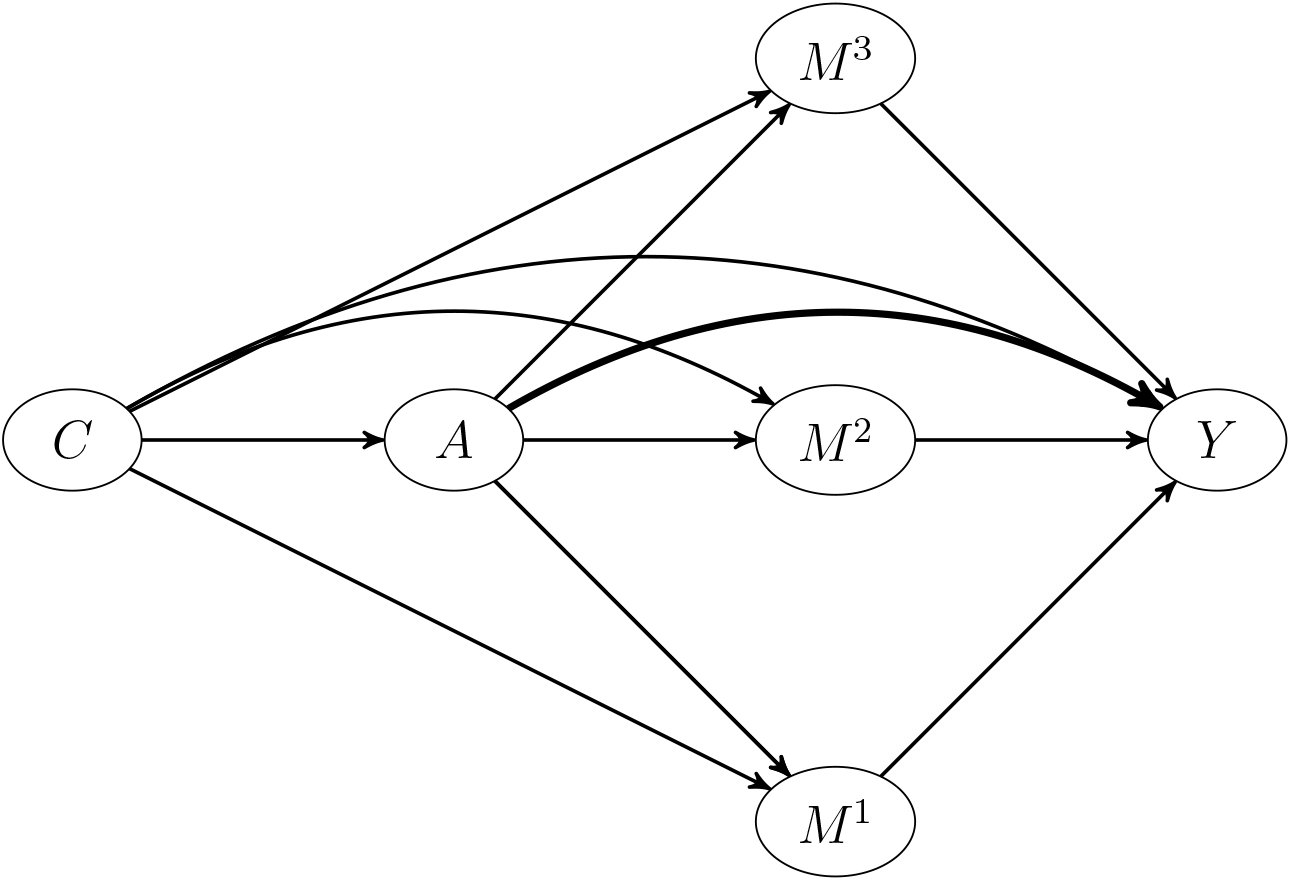
DAG showing causal structure linking exposure, mediators and outcome

The potential outcome setting exposure to *a* and the mediators to *m*^1^,…*m*^*K*^ is denoted 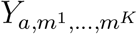. One can also define counterfactuals for each mediator *k*, assuming *A* is set to *a* as 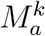. In addition, we will use the following abrieviated notation: 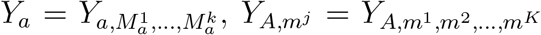 where 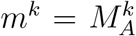 for *k* ≠ *j*, and 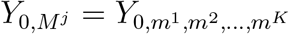, where *m*^*j*^ = *M*^*j*^ and 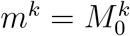 for *k* ≠ *j*.

As is usual with causal inference using the potential outcomes framework, we make Stable Unit Treated Value Assumptions (SUTVA) [18], which implies that the relationship between potential and observed outcomes satisfy ‘consistency’, or that 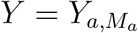 when *A* = *a*. In addition, mediation analysis requires some technical conditional independence assumptions which we will list as needed in the following sections.

### The population attributable fraction

Despite being an intrinsically causal idea, attributable fractions were originally defined via conditional probabilities in a non-causal framework, which has contributed to confusion regarding what they really purport to measure. Here, we will instead use a definiton based on potential outcomes which is now becoming more prominent [9, 8].

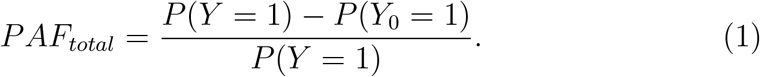

Thinking of *Y*_0_ as the potential outcome for a random individual in a population where no-one was exposed to the risk factor, (1) can be directly interpreted as the relative change in disease prevalence if that risk factor was absent from the population.

### The Pathway Specific population attributable fraction

For a binary risk factor *A* ∈ {0, 1*}*, the PS-PAF for the mediating pathway *A*− > *M*^*j*^− > *Y* is defined as

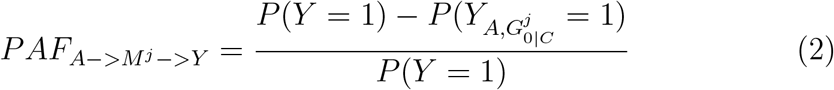

where the random variable 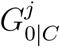 is a random draw from the conditional distribution of 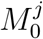 given an individual’s covariates, *C*. Marginally, 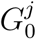 (simulated via first randomly sampling an individual from the population and using their covariates, *C*, to generate 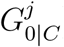) is a draw from the population distribution of 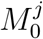, that is the distribution that *M*^*j*^ would have in a hypothetical world where the risk factor was eliminated. Noting this, the PS-PAF can then be interpreted in terms of a population intervention on *M*^*j*^, where its distribution is shifted to the distribution that would be observed in a population where the risk factor *A* was eliminated, with other quantities in the population (covariates *C*, exposure *A* and other mediators *M*^*k*^, *k* ≠ *j*) remaining unchanged. This idea of randomly assigned interventions has previously been introduced to estimate versions of natural direct and indirect effects (sometimes termed interventional direct and indirect effects) in the presence of exposure-induced confounding [**?**]. Comparing PS-PAFs for the various known mediating pathways can help in determining the predominant pathways by which the risk factor is effecting disease in the population. One might also want to compare disease burden attributable to pathways that are either unknown (or involve unobserved mediators). To do this, we will adapt the concept of Direct PAF proposed by Sjölander [13] as follows:

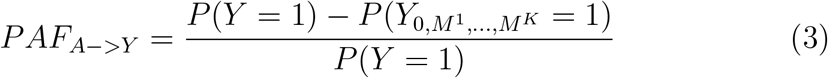

We will refer to equation (3) as the PS-PAF for the direct pathway *A*− > *Y*. (3) measures the disease burden attributable to direct pathogenic pathways from *A* to *Y*, that is any pathway from *A* to *Y* (that may or may not be known) excluding the set of mediating pathways under consideration. The definition in [13] is very similar, except that it refers to the direct pathway with reference to a single mediating pathway, *j*, and will change dependent on that mediating pathway as follows:

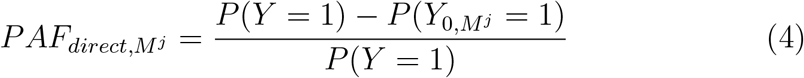

As alluded to in the previous section, 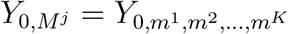, where *m*^*j*^ = *M*^*j*^ and 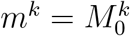 for *k* = *j*, and is not necessarily equal to 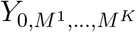.

### Identifiability conditions

Idetifiability conditions are needed to estimate (2) even when values on the exposure, mediators, observed covariates and outcome are available. In particular, conditions 1. and 2. below are necessary to identify PS-PAFs for each mediator *j* of interest.

1. 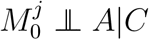 (informally, associations between the risk factor and mediator have causal interpretations within strata of covariates)
2. 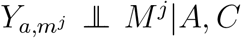, (informally, associations between the mediator and outcome have causal interpretations within joint strata of risk factor and covariates) An additional condition is necessary to identify the *PAF*_*A*−>*Y*_
3. 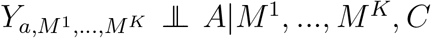, (informally, associations between the risk factor and outcome have causal interpretation within joint strata of mediators (*M* ^1^, …, *M*^*K*^) and covariates)

Under identifiability conditions 1, 2 and 3 we show in the supplementary material that:

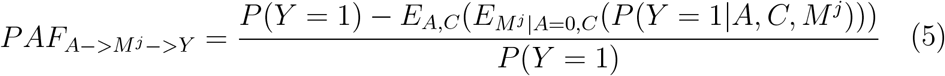

and

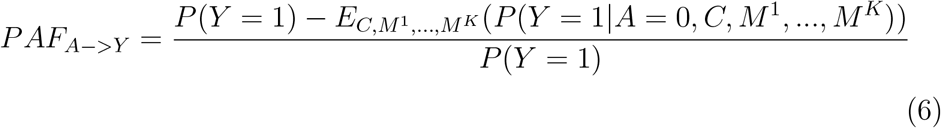

where *E*_*A,C*_(*g*(*A, C*)) = ∫_*a,c*_ *g*(*a, c*)*dF*_*A,C*_(*a, c*) and 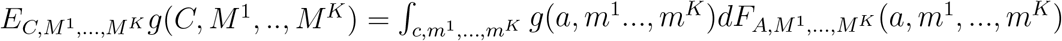 represent expectations in-tegrated over the marginal distributions *F*_*A,C*_ and 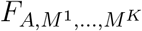of the sub-scripted variables and 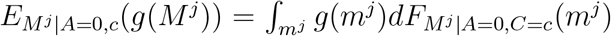 is an expectation integrated according to the conditional distribution of *M*^*j*^ given *A* = 0 and 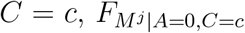. Note that condition 2. implies that there is no post treatment confounding of the mediator outcome relationship; that is there is no variable *L* that is affected by the risk factor *A* such that *L* is a joint cause of *M*^*j*^ and *Y*. If such a variable *L* exists, condition 2. would then become 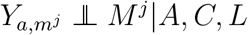, and the pathway specific PAF would change to

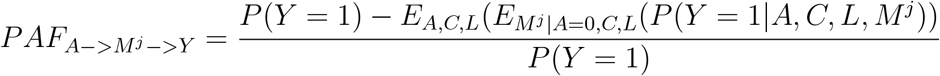

### Mechanistic pathway specific PAF and disabling pathways

Under an extra identifiability condition, the pathway specific PAF can be also expressed in a mechanistic form where the mediator assignment to an individual (within the hypothetical population where the distribution of the mediator is altered) is the mediator that would result for that individual under no exposure to the risk factor:

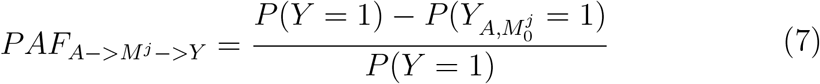

This final condition is:

4. 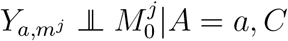

This condition is less intuitive than the conditions 1., 2. and 3. described in the preceding section and involve consideration of cross-world counterfactuals (that is if *a* = 1, 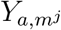 and 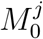 would never be observed on the same individual). However, as shown in the supplementary material, this condition does hold in a non-parametric structural equations model. Under this mechanistic interpretation, the pathway specific PAF can be thought of as the relative change in disease burden in a hypothetical population where the mediated pathway *A*− > *M*^*j*^− > *Y* was disabled. For example, in a simple setting where there is only a single known mediator, *M*, there are 2 potential pathways by which the risk factor effects disease, represented by the pathways *A*− > *M* − > *Y* and *A*− > *Y* in Figure 2(a). The total PAF (which corresponds to a population where a binary risk factor was eliminated corresponds to the disabling both pathways, that is a comparison of disease risk in the populations with causal graphs show in the left hand and right hand panes of Figue 2(a). In contrast, direct PAF involves a comparison of disease risk in the current population with the hypothetical population where the direct pathway is disabled (Figure 2(b)) whereas the pathway specific PAF represents a comparison of the current population with a hypothetical population where the pathway *A*− > *M* − > *Y* is disabled (Figure 2c). Note that in each case, current disease risk is compared to disease risk in some hypothetical population where a pathway has been disabled.

**Figure 2:**
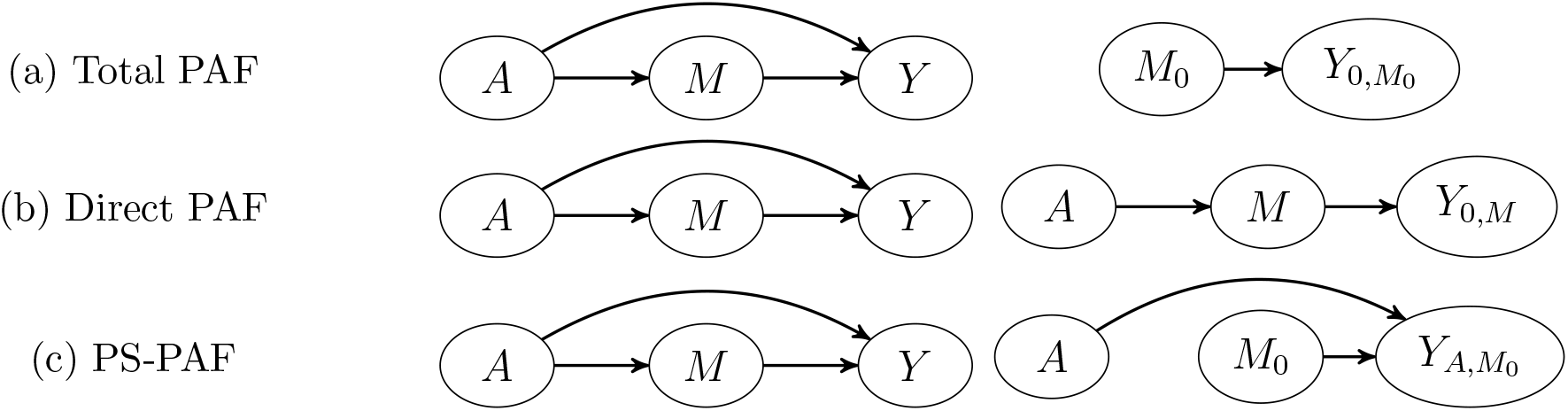
Illustration of total, direct and pathway specific PAF as a comparison between disease risk in current population (graph on LHS) and counterfactual disease risk in a population where particular pathways through which the risk factor effects disease are disabled (graph on RHS). A situation with a single mediated pathway *A*− > *M* − > *Y* is illlustrated. Sub-figure (a) Total PAF compares observed disease risk *P* (*Y* = 1) vs disease risk 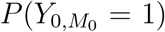 in a hypothetical population where both direct and mediated pathways are disabled. Sub-figure (b) Direct PAF compares observed disease risk vs disease risk *P* (*Y*_0,*M*_ = 1) in a hypothetical population where the direct pathway is disabled. Sub-figure (c) Pathway specific PAF compares observed disease risk Y vs disease risk 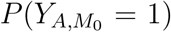 in a hypothetical population where the mediated pathway *A*− > *M* − > *Y* is disabled

### Pathway Specific PAF vs indirect PAF

In addition to proposing the direct PAF (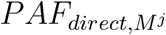, defined in equation (4)) [13] introduced the concept of indirect PAF, defined as:

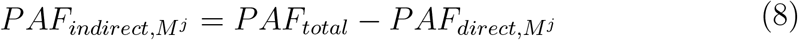

defined in the context of a single mediating pathway, so that the direct and indirect attributable fractions sum to the total PAF. Rather than comparing disease risk in the current and hypothetical populations, (8) implicitly compares disease risk in two hypothetical popualtions: one where the direct pathway *A*− > *Y* has been disabled, with a second where the direct and mediated pathway are both disabled (see Figure 3). Indirect PAF will usually be smaller than the corresponding pathway specific PAF in a one mediator situation, as it is likely that some disease cases in the current population (which are exposed to the risk factor) might be equally well prevented by either eliminating the effect of the direct pathway (that is perhaps 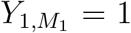, but 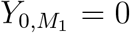, or eliminating the mediated pathway (that is perhaps 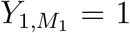, but 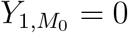). The prevention of such disease cases would contribute to the pathway specific PAF, but not to the indirect PAF.

**Figure 3:**
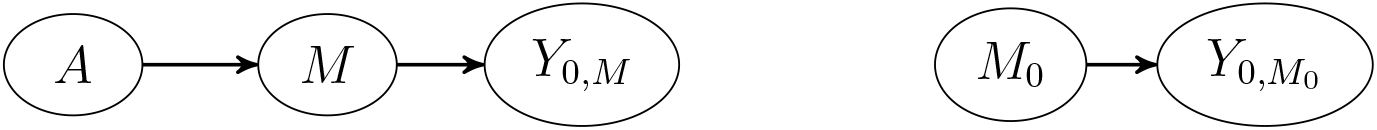
Indirect PAF compares disease risk *P* (*Y*_0, *M*_=1) in a hypothetical population with the direct pathway disabled with the disease risk 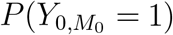 in a hypothetical population with both direct and mediated pathways have been disabled.

### Estimation

In cohort and cross-sectional studies (5) and (6) can be estimated without bias from empirically estimated conditional distributions, given the iden-tifiability conditions discussed earlier. For case control studies, we use a simple-reweighting trick which assumes that the prevalence of disease, *π*, is known and the sampled disease cases and controls are randomly selected from their respective populations. We assume for simplicity that the case:control matching ratio is 1:r, for some *r* ≥ 1. Under these assumptions, the components of (5) and (6) can be found using a re-weighted dataset where cases are assigned weights *w*_*i*_ = 1, and controls are assigned weights *w*_*i*_ = (1*/π* − 1)*/r*. (Note that under the assumptions that prevalence is known and the cases and controls are randomly selected from their source populations, this reweighted sample is a representative sample from the source population; that is means of any variable in this reweighted population agree with the source population, and statistical estimation is consistent). Suppose then, that the researcher specifys and estimates correct models for *P* (*Y*=1 × *A, C*; *M*^1^, …, *M*^*K*^) and a correct conditional model for each *M*^*k*^, conditioned on *A* and *C* (perhaps in the re-weighted population if the original data is from a case-control study). To avoid numerical integration in the continuous mediator case, and simplify the algorithm, we propose that for mediator *j*, 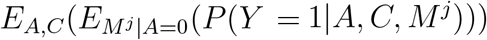 is estimated via the following algorithm. In the algorithm, we set *j* = 1 for simplicity of notation, however an algorithm with obvious modification can be used to estimate the pathway specific PAF for other mediators *j* ≤ *K*. The algorithm assumes that the mediators *M* ^1^, …, *M*^*K*^ are conditionally independent given *A* and *C*, which implies for instance that 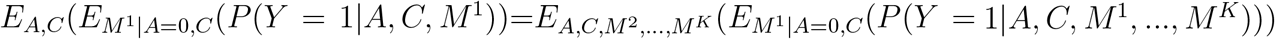. With this assumption, only a single model for the outcome: *P* (*Y* = 1|*A, C, M* ^1^, …, *M*^*K*^) that depends on all mediators *M* ^1^,…,*M*^*K*^ needs to be specified. If the user prefers not to assume this conditional independence condition, the algorithm could be modifed by fitting *K* separate models for Y each using a single mediator; that is the *j*^*th*^ model would regress *Y* on *A, C* and *M*^*j*^ to estimate *P* (*Y* = 1|*A, C, M*^*j*^).

1. Choose a number of simulation iterates, *S*
2. for(j in 1:S){
  a. For each individual in the data, *i*, with exposure, *A*_*i*_, covariate vector *C*_*i*_ and mediators 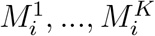 simulate *M* ^1*\**^ from the esti-mated conditional distribution of *M* ^1^ given *A*_*i*_ = 0 and *C*_*i*_
  b. Calculate 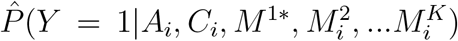 for each *i* using the fitted statistical model
  c. Calculate 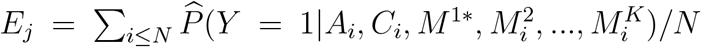 for cohort/cross sectional designs; calculate 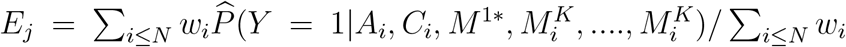 for case control designs.
  }
3. 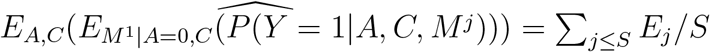

To estimate direct PAF, 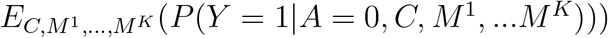 can be estimated more simply by estimating 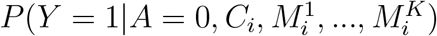 for each individual, and averaging over individuals in the data, taking care to incorporate weighting under a case control design. As an alternative, a double robust estimator for *E* (*P* (*Y*=1 × *A*=0, *C*; *M*^1^, …, *M*^*K*^)] can be derived using the same approaches Sjölander describes in [13].

## Data Example

INTERSTROKE, [20], is a large international case control study designed to quantify the contribution of established risk factors to stroke prevalence at a global level. Here we investigate the possible mediating effects of physical activity on stroke through waist hip ratio, apolipoproteins B to A1 ratio and diagnosis of high blood pressure. We treat waist hip ratio and apolioproteins as continuous variables, whereas diagnosed high blood pressure is binary. Covariates and assumed mediators are as assumed in the causal structure shown by Figure 4. To estimate Sjölander’s direct and indirect attributable fractions, and the pathway specific attributable fractions described above, we fit a main-effects logistic regression predicting stroke status as a function of age,sex,region,eduction diet,physo-social stress factors,smoking status, alcohol use, physical activity, waist hip ratio, apoB/apoA ratio and clinically diagnosed high blood pressure, with the terms for wasit hip ratio and apoB/apoA ratio entering as 5-degree of freedom natural cubic splines. In this regression, stroke controls were upweighted by a factor of 284 to reflect a yearly stroke incidence of first stroke of 0.0035 or 3.5 strokes per 1000 individuals per year, estimated via data from the global burden of disease [21]. To apply the algorithm in the previous section, we also need to simulate from the predicted distribution for the mediators, ApoB/ApoA ratio, waist hip ratio and clincially diagnosed hypertension (conditioned on values for age, sex, education, region, physical activity, diet-score, stress, smoking and alcohol). To do this, we resample residuals from a fitted linear model for ApoB/ApoA ratio and waist hip ratio and add these resampled residuals to the fitted values, whereas for hypertension we simply draw Bernoulli variables with probabilities with probabilities according to the fitted logistic models.

**Figure 4:**
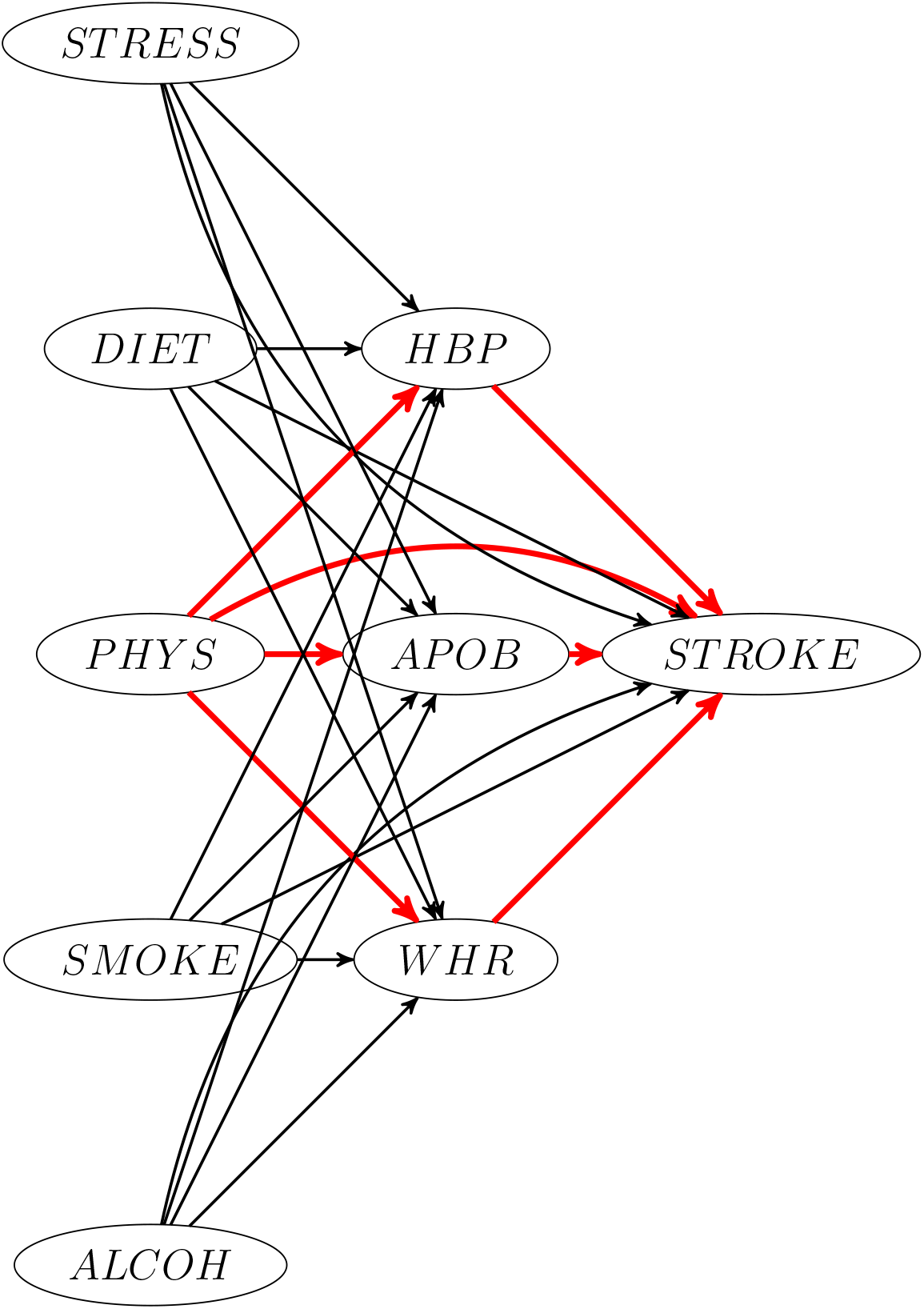
DAG showing assumed causal structure for risk factors in INTERSTROKE. The direct and mediating pathways associated with physical activity are highlighted in bold red. Age, sex and geographic region are confounders for the risk/factor disease relationship for all listed factors on the figure (these are omitted for display purposes)

Note that while there is only one pathway specific PAF from *A* to *Y*, Sjolander’s definition of direct PAF is with reference to a single mediating pathway and will change depending on that pathway. For instance, in this example the direct PAF with reference to high blood pressure essentially pools together all other pathways not going through high-blood pressure. In this example, the mediating pathways and direct pathway due to lack of exercise are likely to increase the probability of stroke, and one would expect PS-PAFs to be somewhat larger than the corresponding indirect PAFs. This pattern is seen in Table 1; the estimated pathway specific PAFs when expressed as percentages range from 3.2%-4.5%, whereas the indirect PAFs range from 0.5% to 1.8%. On the other hand, *PAF*_*A*−>*Y*_ (defined in equation (3)) is 34.3%. According to this analysis, while population disease burden for stroke attributable to physical activity may partially depend on the mediating pathways through blood pressure, waist hip ratio and lipids, it depends heavily on other (unknown) mechanisms. As with any causal analysis, these tentative conclusions depend jointly on correct modeling of conditional probability distributions and on the validity of the causal identifiability assumptions that we listed earlier. Sensitivity analysis [16] could be performed to guage the biases in these results that might be expected due to violations of these conditions. However, the differences between *PAF*_*A*−>*Y*_ and pathway specific mediated PAFs observed here are so large that it would be hard to completely attribute these differences to incorrect modeling or an incomplete causal model.

**Table 1:** Results from estimating the pathway specific, direct and indirect PAFs on the INTERSTROKE dataset using S=100 mediator simulations. Estimated 95% confidence intervals (estimate *±*1.96 *\** standard error) are shown in brackets. Standard errors were estimated using 100 bootstrap iterations, with one bootstrap iteration involving independent resamples from the sample of controls and cases. PAF estimates are rounded to 2 significant digits

| Pathway | total<br>PAF | $PAF_{A \rightarrow Y}$ | $PAF_{Direct, M^j}$ | $PAF_{Indirect, M^j}$ | $PAF_{A \rightarrow M^j \Rightarrow Y}$ |
| --- | --- | --- | --- | --- | --- |
| PHYS->HBP->STROKE | 0.40 (0.34, 0.47) | 0.35 (0.27, 0.42) | 0.39 (0.33, 0.46) | 0.013 (-0.014, 0.039) | 0.041 (0.014, 0.068) |
| PHYS->WHR->STROKE | 0.40 (0.34, 0.47) | 0.35 (0.27, 0.42) | 0.40 (0.33, 0.47) | 0.005 (-0.016, 0.026) | 0.032 (0.007, 0.058) |
| PHYS->APOB->STROKE | 0.40 (0.34, 0.47) | 0.35 (0.274, 0.42) | 0.39 (0.32, 0.45) | 0.018 (-0.005, 0.041) | 0.045 (0.016, 0.073) |

## Discussion

In this paper we have introduced pathway specific attributable fractions as a metric to measure the disease burden attributable to particular exposure mediator pathways. The idea is related to the recently proposed ideas of direct and indirect PAF by Sjölander, but more suitable to multiple mediators [13]. Some differences between the two metrics are summarized in Table 2. In a one mediator situation, one way to understand the distinctions between these concepts is in terms of modified sequential attributable fractions [15], that is attributable fractions that are constructed from disabling pathways in a particular order (as demonstrated in Figures 2 and 3) with the order in which pathways are disabled differing for PS-PAFs and indirect PAFs. In more detail, a PS-PAF can be interpreted as the relative change in disease burden from disabling a particular mediating pathway. The corresponding indirect PAF is also associated with disabling that same mediating pathway, but this time subsequent to disabling the direct pathway. Since the effect of disabling both the direct and mediating pathways is equivalent to the effect of eliminating the risk factor, this effectively forces the additivity property that total PAF is the sum of direct and indirect PAF. Note that in general sequential attributable fractions are constructed to sum to some well-defined overall PAF but usually the sequence corresponds the hypothetical elimination of each of a group of risk factors in some order, rather than disabling pathways for a particular risk factor in some order.

**Table 2:**
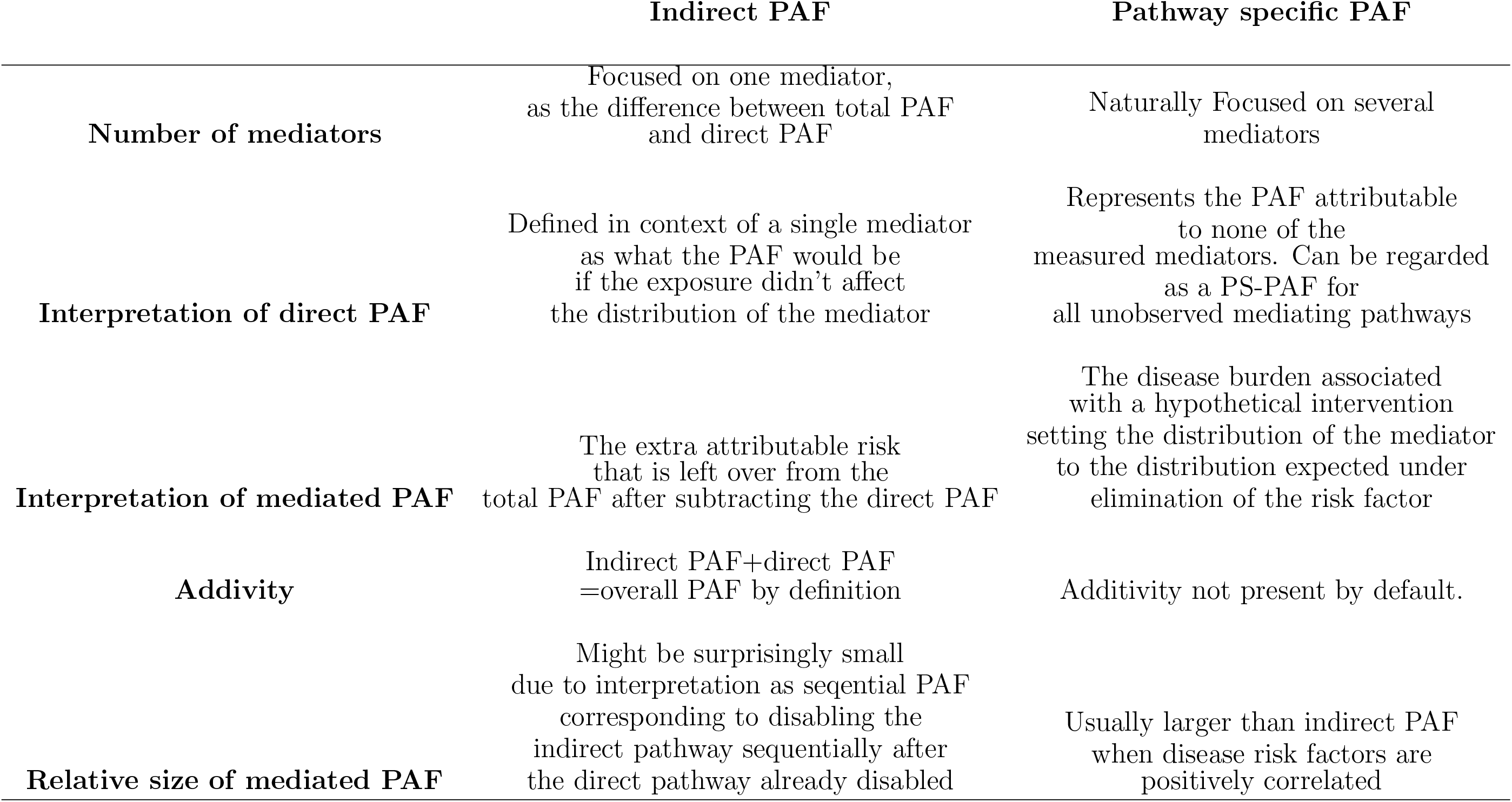
Comparison of PS-PAF with indirect and direct PAF

While this additivity property at first seems appealing, it perhaps is unnatural in the context of attributable fractions, where it is well recognized that the PAF for differing risk factors may sum to more than the joint PAF and sometimes to more than 1 [22]. The sufficient/component cause framework [23] gives a simple but enlightening explanation for this phenomenon. For particular individuals, a certain collection of risk factors (perhaps diet, stress and tobacco usage) might collectively lead to disease at a particular point in time, but the disease may not have occurred at that time if any of the risk factors were not present. This implies that pathway specific PAFs will tend to be larger than indirect PAFs as illustrated in this manuscript, if we view the direct and indirect pathways as independent disease-causing mechanisms.

Whereas attributable fractions can measure the total disease burden associated with a risk factor, there are less useful to measure the real-world impact of a public health intervention on that risk factor since even successful health interventions usually only partially eliminate a risk factor and in addition cannot alter prior history to a risk factor when cumulative exposure might also impact disease. As an example, rather than considering a hypothetical population where smoking is elimated, a realistic population level intervention (such as increasing the tax on cigarettes) may result in a 5% decrease in the number of cigarettes consumed rather than total elimination of smoking. Impact fractions are generalized versions of attributable fractions that measure the reduction in disease prevalence associated with such a population intervention. The ideas described here can easily be adapted to define and estimate pathway specific impact fractions for such real world interventions which may characterize the dominant mechanisms by which the intervention affects disease burden. For example, the pathway specific impact fraction for the pathway *A*− > *M*^*j*^− > *Y* could be defined by letting 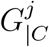 represent a random variable having the population distribution of the mediator *M*^*j*^ under the proposed intervention (simulated again conditional on an individuals covariate vector *C*) and replacing 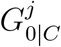 with 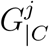 in equation (2).

## Data Availability

R-code for running the analyses in this manuscript can be obtained by contacting the authors

