## supplementary material for "Pathway-specific population attributable fractions"

### Supplementary information

##### Identification of PS-PAF under conditions 1 and 2.

The following is a proof of the identifiability formula for path-specific attributable fractions, using assumptions 1. and 2. detailed in the main manuscript. In the following and subsequent derivations, we assume that covariates, $C$ and mediators $M^1, ...., M^K$ have discrete distributions. The general cases of mixed discrete and continuous random variables with a well defined joint distribution follows similarly by replacing summations with integrals over general probability measures.

$$P(Y_{A,G^j_{0|C}} = 1)$$

$$= \sum_{c,a,m^j} P(Y_{a,G^j_{0|C}} = 1|A = a, C = c, G^j_{0|C} = m^j)P(C = c)P(A = a|C = c)P(G^j_{0|C} = m^j|C = c, A = a)$$

$$= \sum_{c,a,m^j} P(Y_{a,m^j} = 1|A = a, C = c, G^j_{0|C} = m^j)P(C = c)P(A = a|C = c)P(M^j_0 = m^j|C = c)$$

$$= \sum_{c,a,m^j} P(Y_{a,m^j} = 1|A = a, C = c)P(C = c)P(A = a|C = c)P(M^j_0 = m^j|C = c)$$

$$= \sum_{c,a,m^j} P(Y_{a,m^j} = 1|A = a, C = c)P(C = c)P(A = a|C = c)P(M_0^j = m^j|A = 0, C = c)$$

$$= \sum_{c,a,m^j} P(Y_{a,m^j} = 1|A = a, C = c, M = m^j)P(C = c)P(A = a|C = c)P(M_0^j = m^j|A = 0, C = c)$$

$$= \sum_{c,a,m^j} P(Y = 1|A = a, C = c, M = m^j)P(C = c)P(A = a|C = c)P(M = m^j|A = 0, C = c)$$

$$= E_{A,C}(E_{M^j|A=0,C}(P(Y = 1|A, C, M^j)))$$

The first equality follows from the double expectation theorem. The second equality follows from the definition of $G^j_{0|C}$ (which is generated from the distribution of $M_0^j$ conditional on C, independently of A) and the 3rd equality since conditional on $A$ and $C$, $G^j_{0|C}$ is independent of $Y_{a,m^j}$. The fourth equality follows as $M_0^j \perp\!\!\!\perp A|C$. The fifth equality follows since $Y_{a,m} \perp\!\!\!\perp M|A, C$. The sixth equality follows by consistency.

##### Identification of mechanistic PS-PAF under conditions 1, 2 and 4.

$$P(Y_{A,M_0^j} = 1)$$

$$= \sum_{c,a,m^j} P(Y_{a,M_0^j} = 1|A = a, C = c, M_0^j = m^j)P(C = c)P(A = a|C = c)P(M_0^j = m^j|A = a, C = c)$$

$$= \sum_{c,a,m^j} P(Y_{a,m^j} = 1|A = a, C = c, M_0^j = m^j)P(C = c)P(A = a|C = c)P(M_0^j = m^j|A = a, C = c)$$

$$= \sum_{c,a,m^j} P(Y_{a,m^j} = 1|A = a, C = c)P(C = c)P(A = a|C = c)P(M_0^j = m^j|A = a, C = c)$$

$$= \sum_{c,a,m^j} P(Y_{a,m^j} = 1|A = a, C = c)P(C = c)P(A = a|C = c)P(M_0^j = m^j|A = 0, C = c)$$

$$= \sum_{c,a,m^j} P(Y_{a,m^j} = 1|A = a, C = c, M = m^j)P(C = c)P(A = a|C = c)P(M_0^j = m^j|A = 0, C = c)$$

$$= \sum_{c,a,m^j} P(Y=1|A=a,C=c,M=m^j)P(C=c)P(A=a|C=c)P(M=m^j|A=0,C=c)$$

$$= E_{A,C}(E_{M^j|A=0,C}(P(Y=1|A,C,M^j)))$$

Here the proof is almost the same the proof of the path specific PAF. The main difference is the cross world assumption independence assumption: $Y_{a,m^j} \perp\!\!\!\perp M_0^j|A=a,C$ is needed to reduce $P(Y_{a,m^j}=1|A=a,C=c,M_0^j=m^j)$ to $P(Y_{a,m^j}=1|A=a,C=c)$ in the third equality. In contrast, in the previous argument, the equality $P(Y_{a,m^j}=1|A=a,C=c,G^j_{0|C}=m^j)$=$P(Y_{a,m^j}=1|A=a,C=c)$, follows since $G^j_{0|C}$ is randomly generated conditional on $C$ and as a result is independent of $Y_{a,m^j}$ conditional on $A$ and $C$.

##### Identification of $PAF_{A->Y}$ under condition 3.

$$P(Y_{0,M^1,..,M^K}=1)$$

$$= \sum_{c,m^1,...,m^K} P(Y_{0,M^1,...,M^K}=1|C=c,M^1=m^1,...,M^K=m^K)P(C=c)P(M^1=m^1,...,M^K=m^K|C=c)$$

$$= \sum_{c,m^1,...,m^K} P(Y_{0,M^1,...,M^K}=1|C=c,M^1=m^1,...,M^K=m^K,A=0)P(C=c)P(M^1=m^1,..,|C=c)$$

$$= \sum_{c,m^1,...,m^K} P(Y=1|C=c,M^1=m^1,...,M^K=m^K,A=0)P(C=c)P(M^1=m^1,...,M^K=m^K|C=c)$$

$$= E_{C,M^1,...,M^K}(P(Y=1|A=0,C,M^1,...,M^K))$$

The first equality follows from iterated expectation theorem. Here the 3rd identifiability condition: $(Y_{0,M^1,...,M^K} \perp\!\!\!\perp A|M^1,...,M^K$ is used to show $P(Y_{0,M^1,...,M^K}=1|C=c,M^1=m^1,...,M^K=m^K)$=$P(Y_{0,M^1,...,M^K}=1|C=c,M^1=m^1,...,M^K=m^K,A=0)$ in the second equality. The final equality follows from consistency.

##### Non parametric structural equations and the validity of the cross world assumption

Under the assumption that the joint distribution of $(C,A,M,Y)$ follows a non-parametric structural model, the cross world condition: $Y_{a,m} \perp\!\!\!\perp M_0|A=$

$a, C$ is satisfied. For simplicity of notation, we describe a one mediator situation in what follows; a similar argument can be used to demonstrate the same result when there are $K > 1$ mediators. Effectively the non-parametric structural equations model implies the joint distribution is generated sequentially from unknown deterministic functions $F_C, F_A, F_M, F_Y$ as follows:

$$C = F_C(U_C)$$
$$A = F_A(C, U_A)$$
$$M = F_M(C, A, U_M)$$
$$Y = F_Y(C, A, M, U_Y)$$

where, $U_C$, $U_A$, $U_M$ and $U_Y$ are independent noise random variables that add stochasticity to the joint distribution. Potential outcomes can be easily derived using the above equations. For instance, conditioning on A=a and C,

$M_0 = F_M(C, 0, U_M)$ is a function of $U_M$, since $C = F_C(U_C)$ is conditioned on. Also $Y_{a,m} = F_Y(C, a, m, U_Y)$ is a function of $U_Y$, again since $C$ is conditioned on. Since $U_M$ and $U_Y$ are independent, $Y_{a,m} \perp\!\!\!\perp M_0 | A = a, C$
